# Lewis Blood Group Antigens are Associated with Altered Susceptibility to Shigellosis

**DOI:** 10.1101/2020.09.09.20191478

**Authors:** Jhansi L. Leslie, Erin Weddle, Lauren K. Yum, Ye Lin, Matthew L. Jenior, Benjamin Lee, Jennie Z. Ma, Beth D. Kirkpatrick, Uma Nayak, James A. Platts-Mills, Herve F. Agaisse, Rashidul Haque, William A. Petri

**Affiliations:** Department of Medicine, Division of International Health and Infectious Diseases, University of Virginia School of Medicine, Charlottesville, VA, USA; Department of Microbiology, Immunology, and Cancer Biology, University of Virginia School of Medicine, Charlottesville, VA, USA; Department of Statistics, University of Virginia, Charlottesville, VA, USA; Department of Biomedical Engineering, University of Virginia School of Medicine and School of Engineering, Charlottesville, VA, USA; Vaccine Testing Center and Department of Pediatrics, The University of Vermont College of Medicine, Burlington, VA, USA; Department of Public Health Sciences, University of Virginia School of Medicine, Charlottesville, VA, USA; Department of Medicine and Vaccine Testing Center, The University of Vermont College of Medicine, Burlington, VT; Center for Public Health Genomics and Department of Public Health Sciences, University of Virginia School of Medicine, Charlottesville, VA, USA; International Centre for Diarrhoeal Disease Research, Bangladesh, Dhaka

**Keywords:** Blood-group Antigens, Lewis Antigens, Shigellosis

## Abstract

In a cohort of infants, we found that lack of the Lewis histo-blood group antigen was associated with increased susceptibility to shigellosis. Broadly inhibiting fucosylation in epithelial cells *in vitro* decreased invasion by *Shigella flexneri*. These results support a role for fucosylated glycans in susceptibility to shigellosis.

## Introduction

Gram-negative bacteria belonging to the genus *Shigella* are the third leading cause of diarrhea-associated deaths in children under the age of five [1]. The primary treatment for shigellosis is antibiotics, however, globally the incidence of resistance to these drugs is increasing. Additionally, despite the burden of *Shigella* spp. on global health, there are currently no commercially licensed vaccines. Identifying host factors that alter susceptibility to *Shigella* spp. is an important first step in devising novel strategies to protect from this infection.

Blood group antigens are a polymorphic trait that is associated with altered susceptibility to numerous infectious agents including bacteria such as *Helicobacter pylori* and *Campylobacter jejuni*, and viruses [2]. The Lewis blood group antigens are highly expressed in the gastrointestinal tissues making them a plausible target for mucosal pathogens like *Shigella* spp. Lewis antigens are generated via the action of two fucosyltransferases, FUT2/Secretor (α1,2-fucosyltransferase) and FUT3/Lewis (α1, 3/4-fucosyltransferase) [2]. FUT2 modifies a precursor Type 1 oligosaccharide to make the H-antigen, after which FUT3 adds a second fucose to produce the Lewis B antigen. Individuals expressing both enzymes will primarily have Lewis B antigens. While individuals negative for FUT2/Secretor will only express Lewis A antigen. If an individual is FUT3/Lewis negative they will produce neither the Lewis A nor Lewis B glycans.

In a cohort of Bangladeshi infants, we sought to determine if the Lewis histo-blood group antigens were associated with altered susceptibility to shigellosis in the first year of life. We found significant differences in the survival probability free from diarrhea associated with *Shigella* spp. within the first year of life based on inferred Lewis/Secretor genotypes. Using an *in vitro* assay, we showed that broadly inhibiting fucosylation on epithelial cells decreases invasion by *Shigella flexneri*. Together these data suggest a role for fucosylated blood group antigens in susceptibility to shigellosis.

## Methods

### Study Population

We performed a sub-study analysis from the Performance of Rotavirus and Oral Polio Vaccines in Developing Countries (PROVIDE) cohort. The cohort and study design have been previously described [3]. Briefly, 700 infants born in urban Dhaka, Bangladesh were enrolled within the first 7 days of life. During the study, diarrheal episodes were monitored and fecal samples were collected. DNA extracted from the diarrheal fecal samples were used to detect common enteric pathogens using a custom developed TaqMan array cards as described previously [4]. We identified infants with a complete 1-year follow-up, that were successfully phenotyped for Lewis antigens. To exclude subclinical infections, *Shigella* positivity was determined by attributable fraction based on pathogen quantity as previously described [5]. All families participating in the study were consented. PROVIDE was approved by the ethical review boards of the International Center for Diarrhoeal Disease Research, Bangladesh, The University of Virginia, and the University of Vermont. The trial was registered at http://ClinicalTrials.gov (NCT01375647).

### Secretor Status and Lewis Antigen Phenotyping

Lewis and secretor status were inferred from LeA and LeB antigen phenotyping of stored saliva specimens using a dot-blot assay as previously described [6].

### *In vitro S. flexneri* Infections

*S. flexneri* infections were performed as previously described [7] with following modifications. HT-29 cell lines stably expressing yellow fluorescent protein (YFP) membrane marker were maintained in McCoy’s 5A Medium (Gibco) with 10% heat-inactivated fetal bovine serum (Invitrogen). Four days before infection, cells were added to a 96-well cell culture treated plate (Corning). The following day, the medium was replaced with medium supplemented with 250μM 2F-peracetyl-fucose, a cell-permeable inhibitor of fucosyltransferases (2F-PAF, Millipore Sigma), or vehicle (DMSO) [8]. Drug (or vehicle) supplemented medium was replaced each day thereafter, for a total of three days of treatment. Effect of the drug was evaluated by flow cytometry as previously described [8], also see supplemental methods. On the day of infection, the medium was replaced with fresh medium without drug. Cells were infected using a frozen stock made from exponential phase *S. flexneri* serotype 2a 2457T expressing CFP under a β-D thiogalactopyranoside (IPTG)-inducible promoter. Maintenance of virulence plasmid was confirmed by presence of red colonies on plates containing Congo Red. Infection was started by centrifuging the plate for 5 min at 1000 rpm and internalization of the bacteria was allowed to progress for 1 hour before IPTG (10mM final concentration) and gentamicin (50μM) were added to the plate to induce expression of CFP and kill the remaining extracellular bacteria, respectively. Intracellular infection proceeded for another 7 hours at which point cells were fixed with 4% paraformaldehyde and the plates were imaged using the ImageXpress Micro imaging system (Molecular Devices). The number of infection foci per well were manually counted, while infection foci area (size) was measured with ImageXpress imaging software [9].

### Statistical Analysis

Statistical analysis was performed in R version 3.6.3 using standard downloadable packages. Kaplan–Meier (KM) method was used to estimate the probability free from the first *Shigella* attributable diarrheal episode and KM survival plots were generated using the packages survminer, survival, and survMisc. The log-rank test was used to compare the KM survival curves between Lewis and Secretor groups. The Cox proportional hazards model was used to evaluate the risks of *Shigella* attributable diarrhea among Lewis and Secretor groups. A two-tailed Mann-Whitney U non-parametric test was used to compare foci number and area between treatment groups. Data were graphed using the package ggpubR.

## Results

To determine if Lewis or Secretor status is associated with altered susceptibility to shigellosis, we performed a Kaplan-Meier survival analysis. Infants that did not have a diarrheal episode attributable to *Shigella* spp. were censored at one year of age. In this analysis, we found that Lewis histo-blood group status was significantly associated with decreased probability of shigellosis free survival in the first year of life (*P*<0.01), and Le-/Se+ infants were more likely to suffer such illness with a significantly increased hazard ratio of 2.4 (95% CI: *1.4 − 4.2, P=0.001)* compared to Le+/Se+ infants (figure 1A & Supplemental figure1).

**Figure 1:**
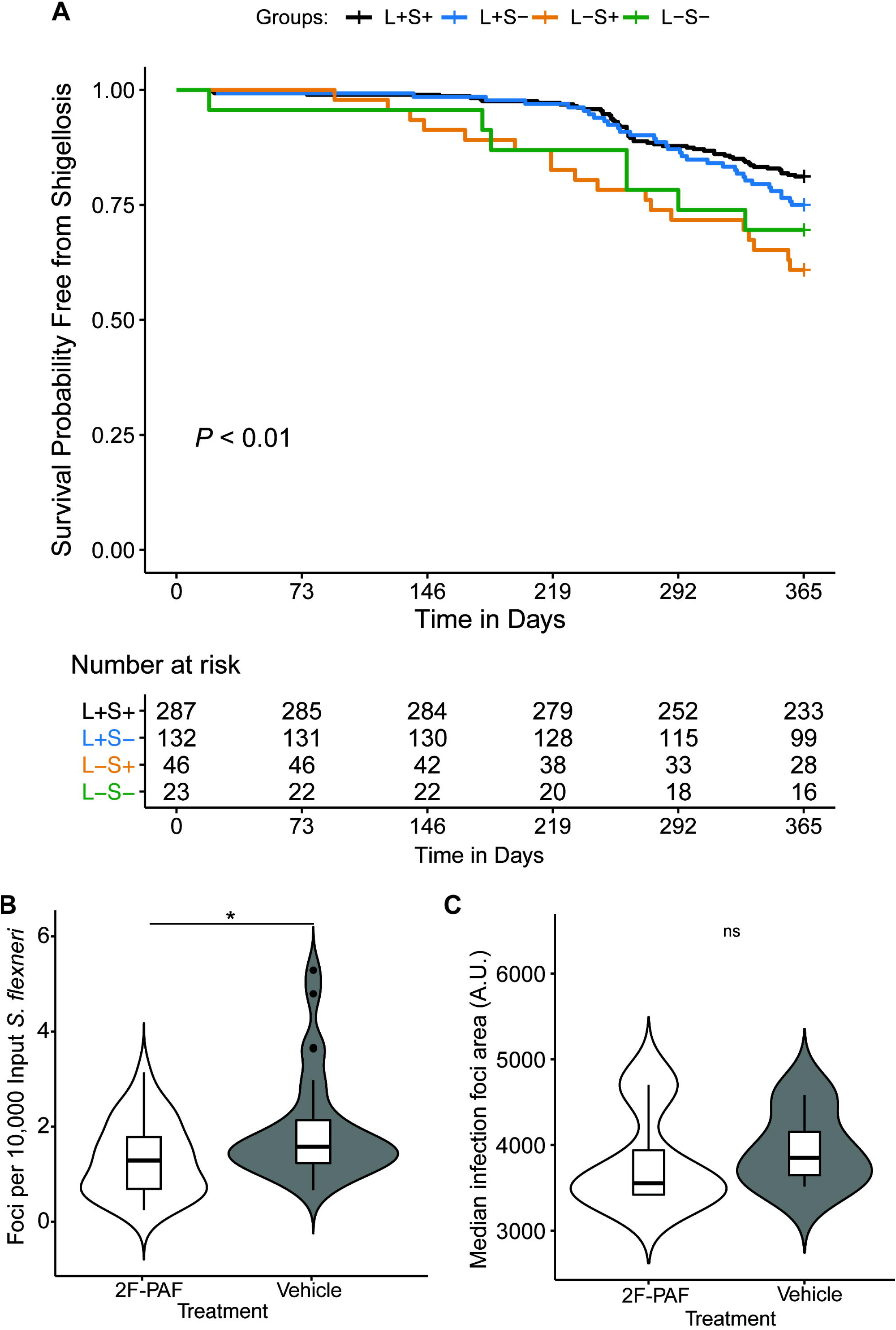
Lewis antigens are associated with altered susceptibility to shigellosis. A) Kaplan-Meier curve showing survival probability free from Shigella spp. attributed diarrheal episode in the first year of life based on inferred Lewis and Secretor status. *P* < 0.01, Log-rank test. Number of children who were at risk for *Shigella spp*. attributed diarrhea are listed at the bottom of the table. B) HT29 cells were treated with 2F-Peracetyl-Fucose (2F-PAF) or vehicle control for three days before infection. 2F-PAF treatment decreased the number of *S. flexneri* infection foci, y axis is infections per well normalized to input inoculum. N = 32 wells per treatment group. Data are combined from four independent experiments. C) 2F-PAF treatment did not alter *S. flexneri* infection foci area (spread). N = 28 wells per treatment group. Data are combined from three independent experiments Significance for both 1B and 1C was determined by a two-tailed nonparametric Mann–Whitney U test. *, *P* < 0.05, ns = not significant.

To test the biological plausibility of this finding, we asked if inhibition of fucosylation in human colonic epithelial cells altered *S. flexneri* infection and/or dissemination. To do this, we utilized a cell-permeable fluorinated fucose derivate 2F-peracetyl-fucose (2F-PAF). Following uptake and metabolism of 2F-PAF into a GDP-fucose memetic, the metabolite broadly inhibits cellular fucoslytransferases. As previously established, treatment of HT-29 cells with 2F-PAF resulted in decreased levels of the fucosylated glycans Lewis A, Lewis B, as well as the Lewis B precursor H-antigen compared to vehicle treated cells (Supplemental figure 2A) [8]. The drug treatment did not alter viability of cells (Supplemental figure 2B). Infection of 2F-PAF treated colonic epithelial cells resulted in significantly decreased numbers of infection foci per well relative to vehicle control (Figure 1B). However, median infection foci area was not significantly different between the treatments (Figure 1C). Together these results suggest that inhibition of fucosyltransferases in colonic epithelial cells decreases *S. flexneri* invasion but does not impact cell-to-cell spread.

## Discussion

In this study, we found that Lewis negative status was associated with increased susceptibility to shigellosis in Bangladeshi infants in their first year of life. This discovery of the role of fucosylation in susceptibility enriches our understanding of shigella pathogenesis and may provide new avenues towards the prevention or treatment of this antimicrobial resistant bacterium.

We demonstrated that inhibition of fucosyltransferases in colonic epithelial cells decreases *S. flexneri* invasion without impacting dissemination. The results of our *in vitro* assay were surprising at first, as one might predict that decreasing fucosylation would mimic the phenotype of the susceptible Lewis-negative infants, resulting in an increase in invasion rather than the observed decrease. However, since the inhibitor used blocks many fucosyltransferases it is possible that our data points to a complex role for other fucosylated glycans in mediating *S. flexneri* invasion of the colonic epithelium. Other groups have found that *S. flexneri* adheres to epithelial cells via glycan-glycan interaction between bacterial LPS and host-cell ABO blood group antigens (which are also fucosylated) in addition to Lewis-antigens [10].

A limitation of the study is that the children were infected with multiple other enteropathogens in addition to *Shigella*. However, the use of attributable fraction rather than solely PCR positivity helped to ascribe a diarrheal infection as shigellosis [5]. An additional limitation was that our detection method targeted the virulence factor *ipaH* which is present in all four species of *Shigella* as well enteroinvasive *E. coli*. *S. flexneri* is one of the most common causes of shigellosis in the region of Bangladesh our cohort was from, thus we used *S. flexneri* for our *in vitro* assays [11].

In addition to bacteria-epithelial cell interactions during infection, Lewis antigens impact neutrophil binding and transepithelial migration [12]. Neutrophil influx is a hallmark of shigellosis, and neutrophils are critical for control of the pathogen [13-15]. While we only tested the effect of fucosylated glycans on *S. flexneri* invasion and dissemination in tissue culture epithelial cells, it is possible that Lewis blood group antigens additionally alter susceptibility to *S. flexneri* via their ability to facilitate recruitment of leukocytes *in vivo*. Together the data we present supports the concept that fucosylated glycans play a role in susceptibility to shigellosis.

## Data Availability

NA

